# Intensified stimulation targeting lateral and medial prefrontal cortices for the treatment of social anxiety disorder: A randomized, double-blind, parallel-group, dose-comparison study

**DOI:** 10.1101/2021.06.08.21258427

**Authors:** Eisa Jafari, Jaber Alizadehgoradel, Fereshteh Pourmohseni Koluri, Ezzatollah Nikoozadehkordmirza, Meysam Refahi, Mina Taherifard, Vahid Nejati, Amir-Homayun Hallajian, Elham Ghanavati, Carmelo M. Vicario, Michael A. Nitsche, Mohammad Ali Salehinejad

**Affiliations:** Department of Psychology, Payame Noor University, Tehran, Iran; Department of Psychology, Faculty of Humanities, University of Zanjan, Zanjan, Iran; Department of Psychology, Mohaghegh-Ardabili University, Ardabil, Iran; Department of Psychology, Shahid Beheshti University, Tehran, Iran; Department of Psychology and Educational Sciences, University of Tehran, Tehran, Iran; Department of Psychology and Neurosciences, Leibniz Research Centre for Working Environment and Human Factors, Dortmund, Germany; Department of Cognitive Science, University of Messina, Messina, Italy; Department of Neurology, University Medical Hospital Bergmannsheil, Bochum, Germany

**Keywords:** Social anxiety disorder, dorsolateral prefrontal cortex (DLPFC), medial prefrontal cortex, VMPFC, transcranial direct current stimulation (tDCS), attentional bias

## Abstract

**Background:** Social Anxiety Disorder (SAD) is the most common anxiety disorder while remains largely untreated. Disturbed amygdala-frontal network functions are central to the pathophysiology of SAD, marked by hypoactivity of the lateral prefrontal cortex (PFC), and hypersensitivity of the medial PFC and amygdala. The objective of this study was to determine whether modulation of dorsolateral and medial PFC activity with a novel intensified stimulation protocol reduces SAD core symptoms, improves treatment-related variables, and reduces attention bias to threatening stimuli.

**Methods:** In this randomized, sham-controlled, double-blind trial, we assessed the efficacy of an intensified stimulation protocol (20 min, twice-daily sessions with 20 min intervals, 5 consecutive days) in two intensities (1 vs 2 mA) compared to sham stimulations. 45 patients with SAD were randomized in three tDCS arms. SAD symptoms, treatment-related variables (worries, depressive state, emotion regulation, quality of life), and attention bias to threatening stimuli (dot-probe paradigm) were assessed before and right after the intervention. SAD symptoms were also assessed at 2-month follow-up.

**Results:** Both 1 mA and 2mA protocols significantly reduced fear/avoidance symptoms, worries and improved, emotion regulation and quality of life after the intervention compared to the sham group. Improving effect of the 2 mA protocol on avoidance symptoms, worries and depressive state was significantly larger than the 1 mA group. Only the 2 mA protocol reduced attention bias to threat-related stimuli, the avoidance symptom at follow-up, and depressive states, as compared to the sham group.

**Conclusions:** Modulation of lateral-medial PFC activity with intensified stimulation can improve cognitive control, motivation and emotion networks in SAD and thereby results in therapeutic effects. These effects can be larger with 2 mA vs 1 mA intensities, though a linear relationship between intensity and efficacy should not be concluded.

## 1 Introduction

With a lifetime prevalence of 13%[1], Social Anxiety Disorder (SAD), also known as social phobia, is the most common anxiety disorder with an early onset age and a risk factor for subsequent depression, substance abuse, and cardiovascular disease[1, 2]. Individuals with SAD *fear* and *avoid* the scrutiny of others and as a result, avoid social or performance situations, and have an intense fear of social situations in which they anticipate being evaluated not only negatively[3] but also positively[4, 5]. SAD is usually associated with other mental health problems particularly elevated anxiety and depressive states[6], decreased quality of life[7], and disturbed emotion regulation[8] leading to disabling consequences such as social isolation and functional impairment. Despite the frequency and severity of the diseases, SAD remains largely undertreated[9].

Exploring new possibilities for the treatment of anxiety disorders, including SAD, through non-pharmacological and/or non-invasive intervention approaches is of growing interest[10-13]. Such an interest arises from an increasing understanding of neurobiological underpinnings of SAD[14] and the difficulties of standard treatment approaches including drug-treatment resistant disorder[15], short-lived efficacy[16] or reluctance to psychotherapy because of being exposed to a feared situation[1], fear of being stigmatized[17], non-affordable expenses, or long waiting list. Recent studies using neuroimaging techniques have highlighted the critical role of cortical and subcortical regions in the pathophysiology of SAD[14, 18-20]. One often-reported finding is hyperactivity of the fear circuit[19]. The amygdala is a key subcortical structure playing an important role in the response to fear and is involved in the pathophysiology of SAD. In patients with SAD, amygdala activation usually increases in response to emotional faces and is correlated with severity of symptoms[2, 21]. In addition to the amygdala, there are other cortical regions with either exaggerated activation such as the medial prefrontal cortex (PFC), anterior cingulate cortex, orbitofrontal cortex[18, 21] or altered activity (e.g. decreased or increased) during emotional and threat processing including dorsolateral PFC (DLPFC)[21-23]. Recently important insights were delivered by nuclear neuroimaging as well. These findings support functional models of SAD by showing abnormalities of neuronal activity in several key limbic and paralimbic regions, including the medial frontal cortex, and also neurochemical pathologies of SAD (i.e., dopaminergic and serotonergic systems)[24].

At the network level, disturbed and aberrant connectivity between the amygdala and frontal cortex[25, 26], and the parietal-occipital network[19] are assumed to be involved in core symptoms of SAD such as emotional reactivity, poor cognitive control over negative emotions and negative self-referential processing[18, 19, 25]. These amygdala-frontal cortex network alterations can be explained as disturbances between the emotion network (involving the amygdala), the motivation network (involving medial and orbitofrontal PFC), and the cognitive control network (involving the DLPFC)[18]. A hyper-responsive emotional network interacts with a sensitive motivation system that rewards and punishes non-social and social stimuli respectively and both of these networks are associated with a diminished cognitive control and emotion regulation network[18]. This is in line with a hot-cold cognition perspective of anxiety disorder where a deficient cold regulatory process (DLPFC, anterior cingulate cortex) and an overresponsive system for emotional and reward/punishment processing (medial PFC, subcortical regions) are involved[20]. Similarly, there are both functional and structural alterations in the parietal-occipital network, and its connections to the anterior network. Here a hyperactivation of medial parietal and occipital regions (posterior cingulate, precuneus, cuneus) and reduced connectivity between parietal and limbic and executive network regions are well-documented[19] that are closely related to negative self-image and biased self-referential processing in patients with SAD. Modulating cortical activity by non-invasive brain stimulation may be useful in order to directly target and alter functionality of the brain networks involved in SAD[10, 27]. Neuromodulation studies that tackle these treatment-relevant variables might therefore be useful for developing innovative treatments for SAD.

Transcranial direct current stimulation (tDCS) is emerged promising for the treatment of anxiety disorders[10-12] and fear response[13]. tDCS is a non-invasive, painless, and well-tolerated brain stimulation technique that applies a weak direct current (typically 0.5 mA– 2 mA) through surface electrodes on the scalp. It can induce acute and neuroplastic alterations of cortical excitability via subthreshold neuronal depolarization and induction of LTP-like plasticity (anodal stimulation), or hyperpolarization and LTD-like plasticity (cathodal stimulation)[28, 29]. tDCS is increasingly used for studying physiological and neurocognitive functions in the healthy brain, and for clinical applications (for a detailed review see[30, 31]). Cognitive, emotional and motivational functions of the lateral and medial PFC are increasingly being studied with tDCS[32-36]. The efficacy of tDCS, especially for therapeutic purposes, depends on different factors including stimulation parameters (e.g. target region, intensity, duration, repetition rate, repetition interval)[37-39] some of which such as target region, repetition rate and interval, are not well-studied in randomized clinical trials in SAD. Very few studies so far, have investigated efficacy of tDCS in SAD. A recent tDCS study investigated the effects of single-session anodal DLPFC stimulation on attentional bias to threat in SAD and found a significant decrease of attentional bias during anodal tDCS[40]. No other tDCS study in SAD is available at present. Two case studies, that applied low-frequency excitability-diminishing repetitive transcranial magnetic stimulation (r-TMS) over the right ventromedial PFC (VMPFC) of three patients with SAD resulted however in a significant reduction of anxiety levels after 12 sessions of rTMS[41, 42].

In order to evaluate the potential of tDCS for the treatment of SAD, trials with longer courses of stimulation, inclusion of primary and secondary clinical scales and cognitive measures, and monitoring of long-term outcomes are required. Accordingly, in this registered, randomized, sham-control clinical trial we aimed to (1) investigate the effects of intensified tDCS (repeated stimulation with 20 min interval) over the left DLPFC and medial PFC on primary and secondary clinical variables in patients with SAD, (2) compare the effect of different stimulation intensities (1 vs 2 mA) on treatment efficacy in comparison to the sham condition, (3) examine the effects of these interventions on attentional bias to threat in patients with SAD, and (4) to see whether the expected improvement in attentional bias to threat is correlated with clinical improvement and symptom reduction. The rationale behind this stimulation protocol is related to functional *relevance* of target regions and stimulation *parameters*. With regard to the former, our protocol was designed to upregulate the executive control network via anodal DLPFC stimulation and downregulate exaggerated emotional reactivity by stimulation of the medial PFC via its connection to subcortical limbic structures[10, 18]. With regard to stimulation parameters, we were interested to examine the efficacy not only of different stimulation intensities but also of repetitive stimulation with a short (20 min) interval, which induces late-phase plasticity at the physiological level[43]. This is the first tDCS study in SAD with a randomized parallel-group design that explores the effects of a novel intensified tDCS intervention at two different stimulation intensities on symptoms reduction and attentional bias.

## 2. Materials and methods

### 2.1. Participants

Fifty-six individuals with SAD (18-50 years, mean age=32.36±6.99) were initially recruited from those referred to six Neuropsychiatric Clinics in Ardabil, Iran. Forty-five patients who met the inclusion criteria were randomized into three study arms. All patients were diagnosed with SAD according to the DSM-5 criteria[3] by experienced, licensed psychiatrists. Sample size was a-priori calculated (f=0.35 equivalent to a medium partial eta squared of 0.10, α=0.05, power=0.95, *N*=30, for a mixed-model ANOVA with 3 measurements, group (1 mA, 2 mA, sham) as the between-subject factor and time (pre, post, follow-up) as the within-subject factor. We added 15 more participants to compensate for potential dropouts. Four patients from all groups could not complete the whole treatment, and thus final analysis of post-intervention measurements was conducted on 41 participants (1mA tDCS *N*=14, 2mA tDCS *N*=13, sham tDCS *N*=14). The follow-up measurement was conducted in 36 patients (12 subjects per group) due to dropout of five more participants (Fig 1). The inclusion criteria were: (1) diagnosis of SAD via the structured clinical interview according to the DSM-5, (2) 18-50 years old, (3) non-smoker, (4) no previous history of neurological diseases, brain surgery, epilepsy, seizures, brain damage, head injury or metal brain implants, and (5) absence of other psychiatric disorders except for SAD, as confirmed by a structured clinical interview conducted by a professional licensed psychiatrist. Overall, 19 patients had comorbidities with other anxiety disorders, including generalized anxiety disorder, post-traumatic stress disorder, and panic disorder (Table 1). Patients who were taking anxiolytic (BZD; n=23) and/or anti-depressant (SSRI; n=12) medication had a stable medication regime from one month before the start of the study up to the follow-up. All participants were native speakers and had normal or corrected-to-normal vision. The study was a registered clinical trial (trial ID: [IRCT20181013041327N2](https://en.irct.ir/trial/44041)) and performed according to the Declaration of Helsinki, approved by the Ardabil University of Medical Science Ethics Committee (Ethics code: IR.ARUMS.REC.1398.478). Participants gave their written informed consent before participation. See Table 1 for demographics.

**Table 1.** Demographic data

|  |  | 1 mA tDCS | 2 mA tDCS | sham tDCS | *p*-value* |
| --- | --- | --- | --- | --- | --- |
| Sample size (*n*) |  | 14 | 13 | 14 |  |
| Age – Mean (SD) |  | 32.83 (7.46) | 33.66 (6.19) | 30.58 (7.46) | 0.611 |
| Sex – Male (female) |  | 6 (6) | 5 (7) | 7 (5) | 0.894 |
| Marital Status – Single (married) |  | 6 (6) | 5 (7) | 7 (5) | 0.717 |
| Duration of disease- mean (SD) |  | 4 (1.85) | 3.50 (1.78) | 4.16 (2.24) | 0.370 |
| On medication (*n*)* |  | 7 | 7 | 9 | 0.890 |
| Type of medication | BZD | 3 | 4 | 4 | 0.837 |
|  | SSRI+BZD | 4 | 3 | 5 |  |
| Comorbid other anxiety disorder | GAD | 3 | 3 | 2 | 0.671 |
|  | Panic attacks | 3 | 2 | 3 |  |
|  | PTSD | 0 | 2 | 1 |  |
| Education | Diploma & higher | 7 | 8 | 7 | 0.890 |
|  | Under diploma | 5 | 4 | 5 |  |

*Note:* tDCS = transcranial Direct Current Stimulation; M = Mean; SD = Standard Deviation; BZD = Benzodiazepines; SSRI = selective serotonin reuptake inhibitor; GAD = generalized Anxiety Disorder; PTSD = Post-traumatic Stress Disorder; \* = between group differences in demographic variables were explored by Chi-square tests or Fisher's exact test for categorical variables and F-tests for continuous variables. \* = medication included anti-anxiety alone (*n* = 11) and anti-anxiety + anti-depressants (*n* = 12).

**Fig. 1:**
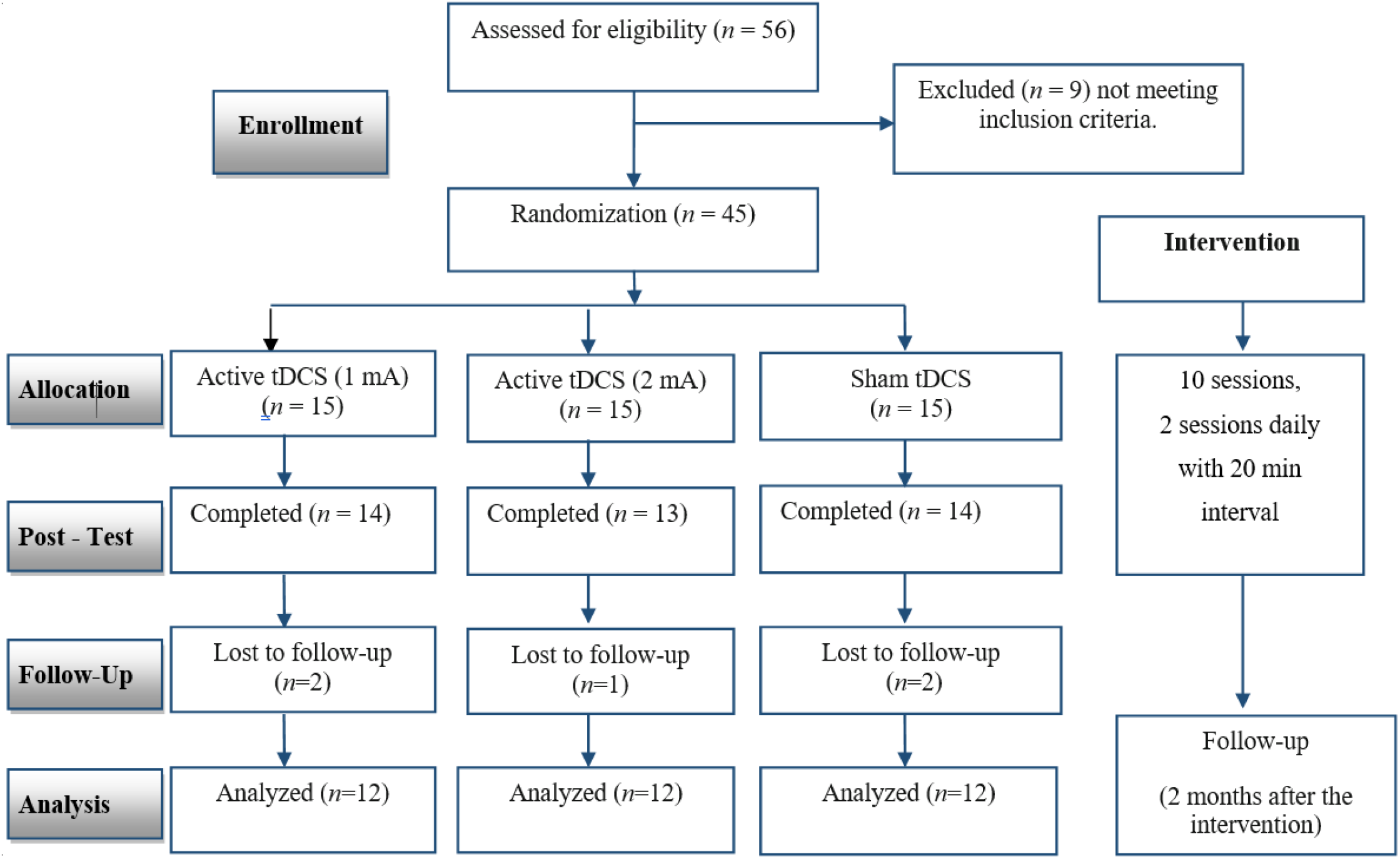
CONSORT Flow Diagram of study inclusion. 41 patients completed the post-intervention measurement, and 36 patients completed the follow-up measurement of the primary clinical symptoms during the lockdown.

### 2.2. Measures

#### 2.2.1. Primary measure: Liebowitz Social Anxiety Scale (LSAS)

The LSAS is one of the most commonly used clinical scales for the assessment of social phobia[44]. It is a 24-item scale assessing the range of social interaction and performance situations that individuals with SAD may *fear* and/or *avoid*, and a valid and treatment-sensitive measure of social phobia[44]. The clinician asks patients to rate *fear* and *avoidance* during the past week on a 0–3 Likert-type scale with 0 and 3 indicative of “never” (0%) and “usually” (67%-100% of the time) occurring. The overall total score is calculated by summing up fear and avoidance scores. The LSAS has a Cronbach’s alpha of 0.96 for the overall score and 0.92 for total fear and total avoidance scores, respectively[44]. Cronbach’s alpha in our sample was 0.91 for the overall score, 0.90 for the “fear” subscale, and 0.92 for the “avoidance” subscale. Patients’ symptoms were evaluated before (pre-intervention), immediately after (post-intervention), and two months after the end of the intervention (follow-up).

#### 2.2.2. Secondary measures: worry, depressive state, emotion regulation and quality of life

Additionally, we measured worry with the Penn State Worry Questionnaire (PSWQ)[45], depressive states with the Beck Depression Inventory-II (BDI-II), emotion regulation with the Difficulties in Emotion Regulation Scale (DERS)[46], and quality of life with the WHOQUL questionnaire[47]. These measures were used to assess the clinical efficacy of the intervention on additional aspects impaired in patients with SAD[7, 48, 49]. Patients completed all measures before and after the intervention. A detailed description of these measures is presented in the supplementary materials.

#### 2.2.3. Attentional bias task

In addition to clinical measures, attention bias to threat stimuli was measured using the dot-probe paradigm[50]. Attention bias is considered a robust cognitive construct for measuring treatment efficacy in emotional disorders[51]. Studies using this paradigm have shown that social anxiety is associated with an attentional bias towards disorder-related stimuli such as threatening faces[52]. Two stimuli are presented to the participants that appear randomly on either side of the screen for a pre-determined time before an asterisk is presented at the location of one former stimulus. Participants are instructed to indicate the location of the asterisk as quickly as possible via the response box. In this computerized task, 80 pictures of facial emotional expressions (40 males, 40 females) were selected from the Warsaw Set of Emotional Facial Expression Pictures[53]. 40 pictures represented threat-and-fear-related facial expressions and 40 ones represented neutral facial expressions. Stimuli of each condition (threat-related vs neutral) were presented in each measurement in random order. During the task, a fixation cross was shown initially in the center of the screen for 1000 ms. A pair of facial pictures (threat/neutral, neutral/neutral) were then presented simultaneously on the left and right side of the screen from 500 ms. Immediately following the presentation of faces, a dot probe was presented at the location of one of the faces (threat vs neutral). Participants were asked to indicate the location of the probe by pressing the corresponding key as fast as possible. Their reaction time (RT), as a measure of selective attention, and bias score (mean difference between threat-related stimuli – neutral stimuli) were calculated as the primary outcome measure. Details about stimuli presentation are displayed in Fig 2B. Stimuli presentation was controlled by a laptop with a 15.2” screen[54], at a viewing distance of approximately 50 cm.

**Fig. 2.**
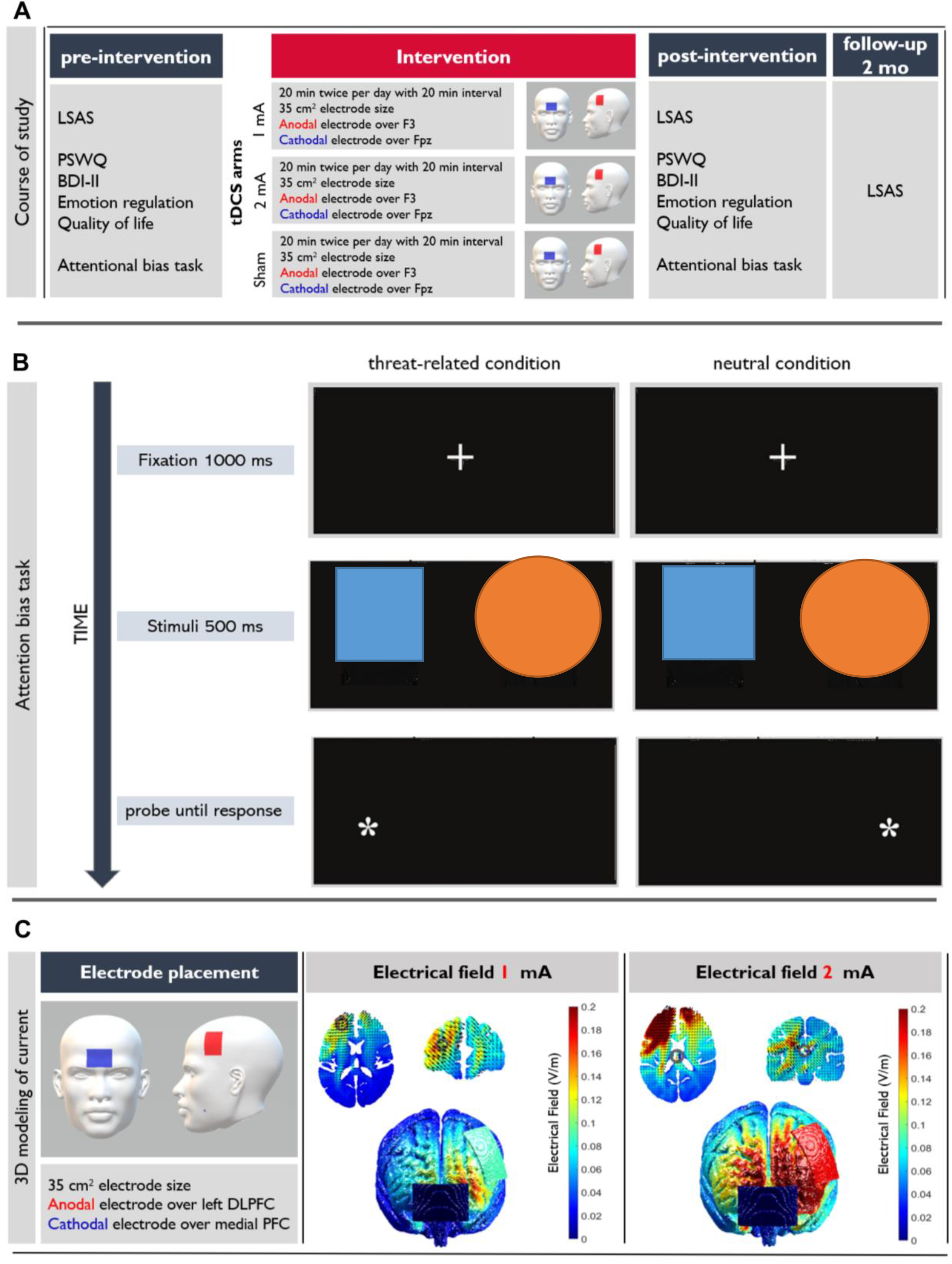
The course of the study (A), attentional bias task characteristics (B), and 3D modeling of the electrical current flow in the head (C) [Legend at the end] *Note*: in B photos are removed to meet the guidelines the pre-print server.

### 2.3. tDCS

Direct currents were generated by an electrical stimulator (NeuroStim 2, Medina Teb, Iran) and applied through a pair of saline-soaked sponge electrodes (7×5 cm) for 20 min with 15 s ramp up and 15 s ramp down. In both active (1 mA, 2 mA) and sham conditions, anodal and cathodal electrodes were placed over the left DLPFC (F3) and medial PFC (Fpz) respectively, according to the 10–20 International EEG System. For sham stimulation, electrical current was ramped up for 30 seconds to generate the same sensation as in the active condition and then turned off without the participants’ knowledge[55, 56]. The two experimenters who applied tDCS were blind to study hypotheses but not to the tDCS conditions and did not participate in any other parts of the study. A side-effect survey was done after each tDCS session (Fig. 2a). A 3D model of the current flow in the head was created to determine induced electrical fields in the brain for the above-mentioned tDCS protocol (anodal F3 - cathodal medial PFC) using ROAST [57], an open-source pipeline for transcranial electrical stimulation (tES) modeling. Details of the modeling procedure are summarized in Fig 2C and supplementary information.

### 2.4. Procedure

Prior to the experiment, participants completed a brief questionnaire to evaluate their suitability for brain stimulation. All groups of participants received 10 sessions of stimulation (2 sessions daily, 1 week in total) with 20 min intervals between daily sessions. Participants were asked to sit relaxed in a waiting room during stimulation intervals. Clinical measures (e.g. social anxiety symptoms, worry, depressive states, emotion regulation and quality of life) and attention bias task performance were evaluated immediately before the first tDCS session (pre-intervention), right after the end of the last tDCS session (post-intervention), and 2 months following the last stimulation session (follow-up) (Fig 2A). tDCS sessions were conducted between 2:00-5:00 PM in all patients and across all sessions. The 2-month follow-up measurement was conducted during the COVID-19 pandemic and participants were not allowed to physically attend the assessment session. We did, however, assess the LSAS, as the primary clinical measure, via an online interview. Patients were instructed about the task before the beginning of the experiment. Except for the follow-up session which took place remotely, each measurement session took around 2 hours. To guarantee double-blinding, examining outcome measures, data analysis and group assignment were performed by independent researchers who were not involved in delivering stimulation sessions.

### 2.5. Study design and statistical analysis

Our study had a randomized, double-blind, parallel-group design to prevent blinding failure and carry-over effects. Participants were blind to study hypotheses and stimulation conditions. The experimenter who conducted the outcome measures was blinded to the tDCS conditions. To guarantee blinding of this investigator, tDCS was applied by other investigators[56]. Data analyses were conducted with the statistical package SPSS, version 27.0 (IBM, SPSS, Inc., Chicago, IL). The normality and homogeneity of data variance were confirmed by Shapiro-Wilk and Levin tests, respectively. Mixed model ANOVAs were conducted for the dependent variables (LSAS, PSWQ, BDI-II, WHOQUL scores; attention bias task RT) with “group” (active 1mA, active 2mA, sham) as the between-subject and time (pre-intervention, post-intervention, follow-up only for LSAS) as the within-subject factors. Mauchly’s test was used to evaluate the sphericity of the data before performing the respective ANOVAs (*p*<0.05). In case of violation, degrees of freedom were corrected using Greenhouse-Geisser estimates of sphericity. Post *hoc* analyses were calculated using Bonferroni-corrected Student’s t-tests and included the pairwise comparisons of interest within intervention groups (pre-intervention vs post-intervention, pre-intervention vs follow-up for LSAS only) and between intervention groups (pre-intervention: 1 mA vs 2 mA; post-intervention: 1 mA vs 2 mA). Associations between clinical symptoms and behavioral performance were explored via the Pearson correlation (*p*<0.05).

## 3. Results

### 3.1. Data overview

Patients tolerated the stimulation well and no adverse effects were reported during and after stimulation. No significant difference was found between the group ratings of tDCS side effects (Table S1). The mean and standard deviation of the dependent variables before and after the intervention is presented in Table 2. No significant between-group differences were observed for baseline measurements.

**Table 2:**
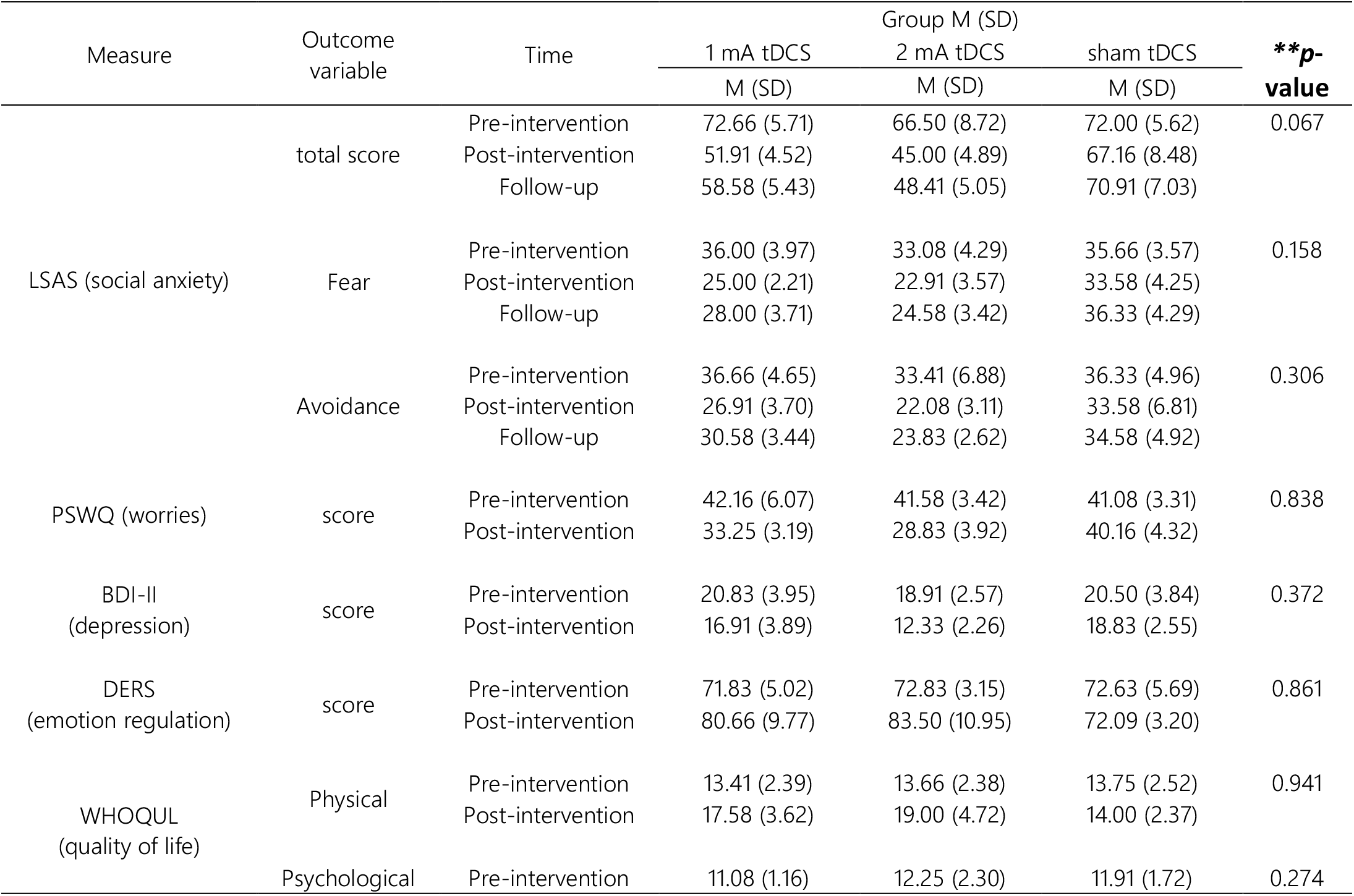
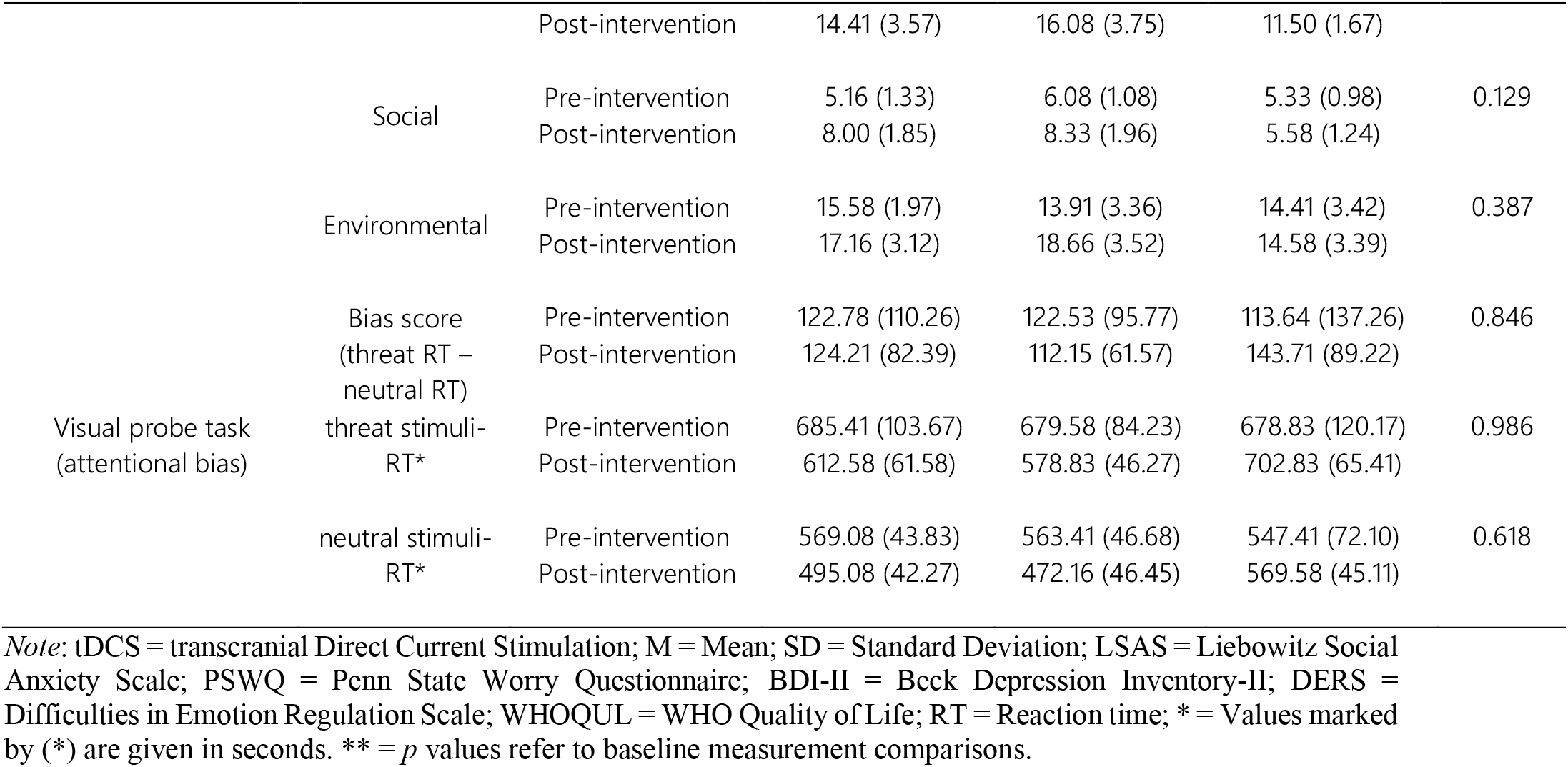
Means and SDs of social anxiety symptoms, clinical measures, and attentional bias task performance before and after the intensified tDCS interventions.

### 3.2. Efficacy of tDCS on SAD symptoms

The results of a 2 (domain: fear, avoidance) ×3 (time: pre, post, follow-up) ×3 (group) mixed ANOVA revealed a significant interaction of group×time (*F*_*2.29,66*_=10.7, *p*<0.001, *η*p2=0.37) on LSAS scores. The interaction of group×time×domain, group×domain, and time×domain was not significant (Table 3) indicating that fear and avoidance subscales were not differently affected by the intervention. Bonferroni-corrected post *hoc* analyses (*p*=0.005) showed a significant decrease of both *fear* and *avoidance* scores at post-intervention and 2-month follow-up measurements in both, 1 mA (post-intervention: *t*_*fear*_=7.18, df=35, *p*<0.001; *t*_*avoidance*_=4.98, df=35, *p*<0.001; follow-up: *t*_*fear*_=5.22, df=35, *p*<0.001; *t*_*avoidance*_=3.11, df=35, *p*=0.003) and 2 mA groups (post-intervention: *t*_*fear*_=6.64, df=35, *p*<0.001; *t*_*avoidance*_=5.79, df=35, *p*<0.001; follow-up: *t*_*fear*_=5.55, df=35, *p*<0.001; *t*_*avoidance*_=4.90, df=35, *p*<0.001) compared to pre-intervention, but no significant alterations in the sham group (*t*_*fear*_=1.35, df=35, *p*=0.531; *t*_*fear*_=0.43, df=35, *p*>0.999; *t*_*avoidance*_=1.43, df=35, *p*=0.487; *t*_*avoidance*_=0.89, df=35, *p*>0.999). When compared to the sham group, both interventions significantly reduced *fear* and *avoidance* after the intervention (1 mA: *t*_*fear*_=5.60, df=35, *p*<0.001; *t*_*avoidance*_=3.43, df=35, *p*=0.002; 2 mA: *t*_*fear*_=6.97, df=35, *p*<0.001; *t*_*avoidance*_=5.88, df=35, *p*<0.001). *Fear* symptom improvement continued up to 2 months after the intervention in both groups but the *avoidance* symptom showed significant improvement in the follow-up only in the 2 mA group (1 mA: *t*_*fear*_=5.44, df=35, *p*<0.001; *t*_*avoidance*_=2.04, df=35, *p*=0.129; 2 mA: *t*_*fear*_=7.67, df=35, *p*<0.001; *t*_*avoidance*_=5.51, df=35, *p*<0.001) (Fig 3a,b). Baseline between-group comparisons (active groups vs sham) showed no significant differences pre-intervention, indicating that reduced *fear* and *avoidance* symptoms were specific to tDCS effects. No significant difference was found between intervention groups (1 vs 2 mA) for the fear scores in the post-intervention (*p*=0.524) and follow-up (*p*=0.083) measurement. The reduction of the avoidance symptom was, however, significantly larger in the 2 mA vs 1 mA condition right after (*t*=2.47, df=35, *p*=0.045) and 2 months (*t*=3.45, df=35, *p*=0.002) following intervention (Fig. 3a,b). Individual scores of LSAS, as the primary measure, are displayed in scatter plots for each group in the supplementary materials (Fig. S1).

**Table 3:**
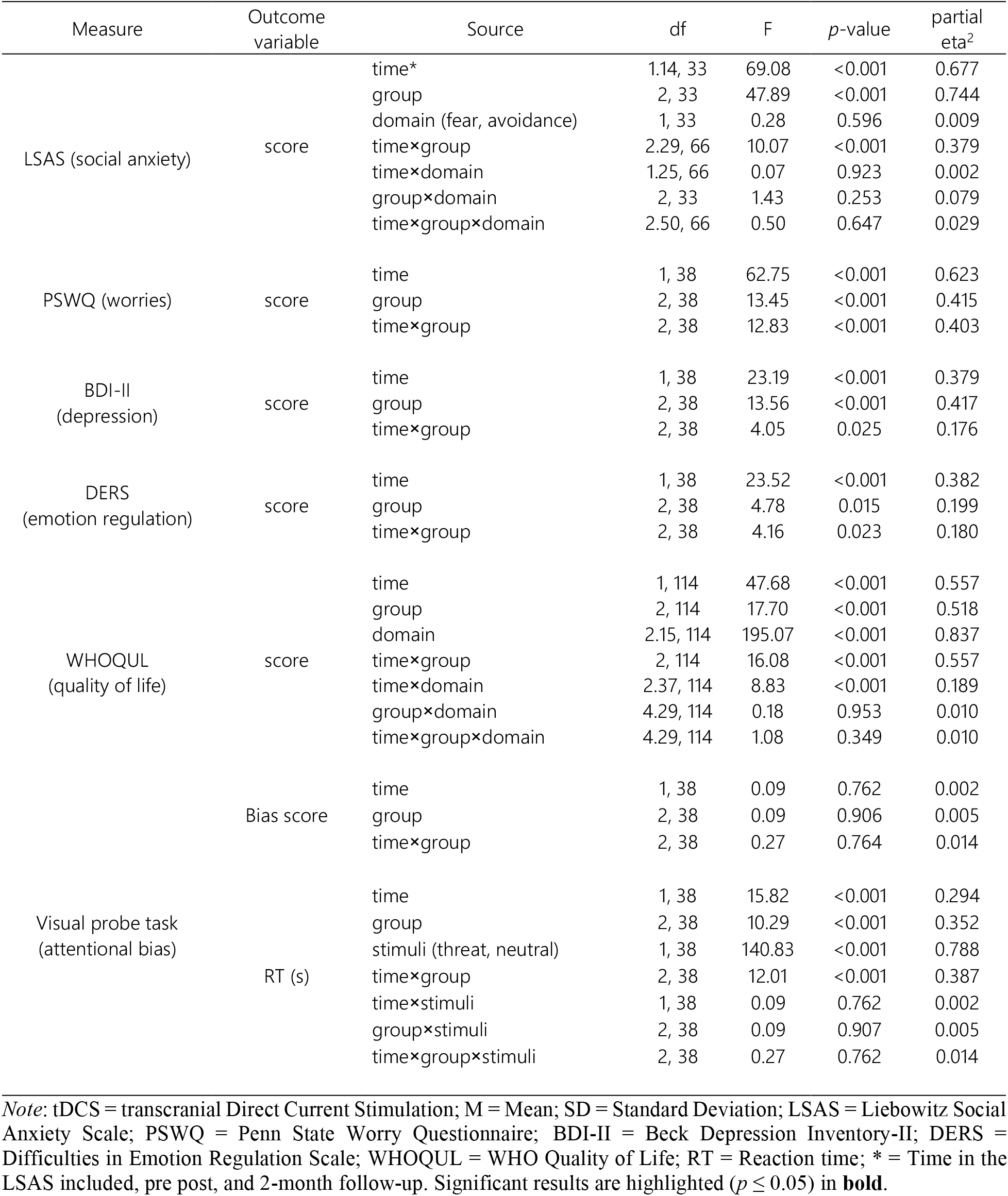
Results of the mixed model ANOVAs for effects of group (1 mA, 2 mA, sham) and time (pre-intervention, post-intervention) on social anxiety symptoms, clinical measures, and attentional bias task performance.

**Fig. 3:**
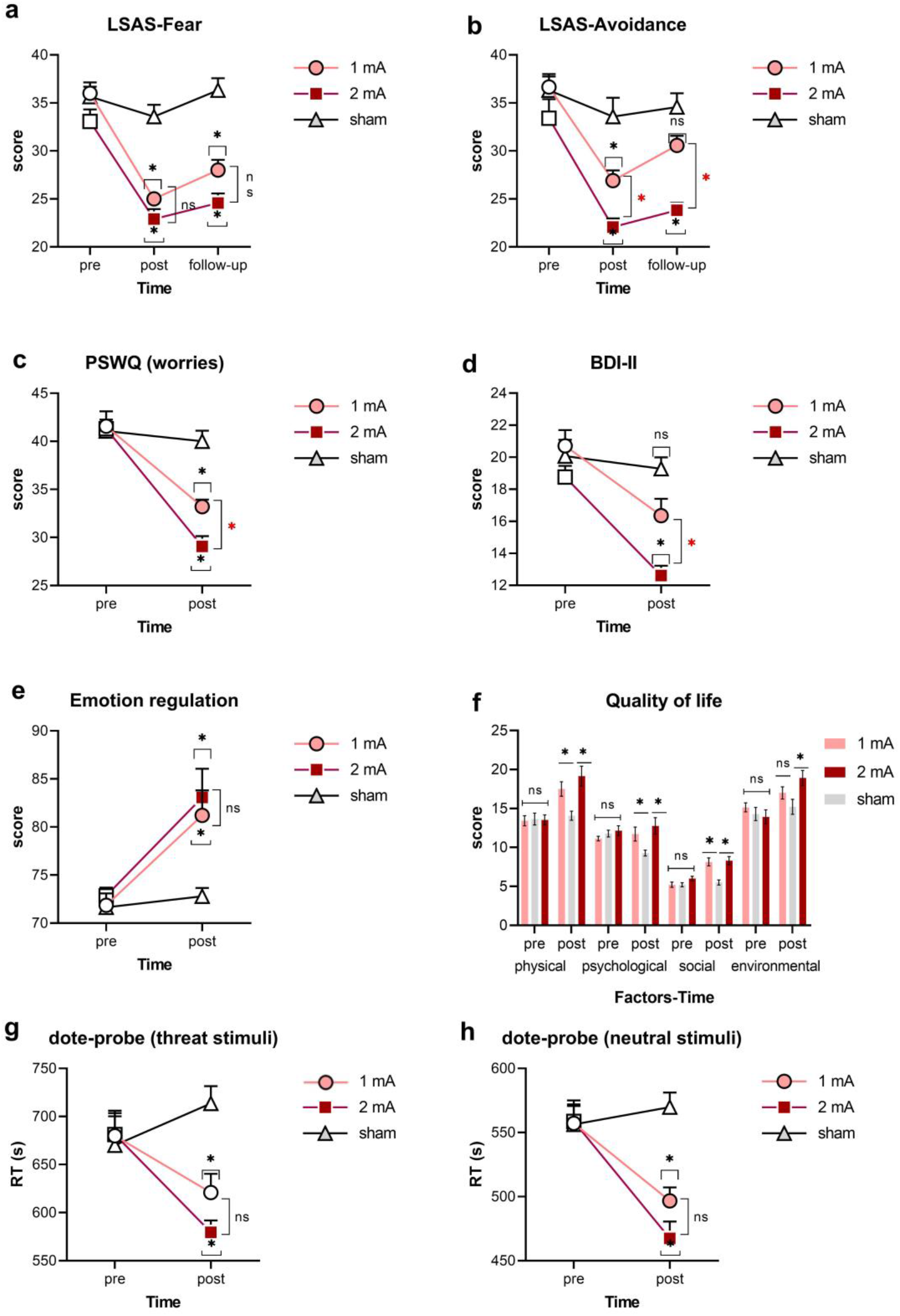
The effect of the intervention on SAD symptoms, treatment-related variables and attention bias to threat. SAD symptoms measured by LSAS (a,b), treatment-related variables (c-f) (worries measured by PSWQ, depressive state measured by BDI-II, emotion regulation measured by DERS and quality of life measured by the WHO quality of life scale), and attentional bias to threat-related and neutral stimuli (g,h) before and immediately after intervention. The SAD symptoms were measured at the 2-month follow-up as well remotely (due to pandemic situation). *Note*: LSAS = Liebowitz Social Anxiety Scale; PSWQ = Penn State Worry Questionnaire; BDI-II = Beck Depression Inventory-II; DERS = Difficulties in Emotion Regulation Scale; WHOQOL = WHO Quality of Life scale; RT = Reaction time; s = seconds; Filled symbols represent a significant difference between pre-intervention measurement vs post-intervention and follow-up measurements (if available) within intervention groups. Floating asterisks [*] indicate a significant difference between active stimulation groups (1 and 2 mA) vs sham tDCS at a specific time point. Asterisks next to the brackets indicate a significant difference between active tDCS groups at a specific time point. ns = nonsignificant. All pairwise comparisons were conducted using Bonferroni corrected t-tests. All error bars represent s.e.m.

### 3.3. Efficacy of tDCS on SAD secondary clinical measures

For worries, the results of the 2×3 mixed model ANOVA showed a significant group×time interaction (*F*_*2,38*_=12.83, *p*<0.001, *η*p2=0.40) on PSWQ scores. The main effects of group and time were significant as well (Table 3). Bonferroni-corrected post *hoc* t-tests (*p*=0.016) showed a significant decrease of worries between pre- and post-intervention measurements in the 1 mA (*t*_*40*_=5.56, *p*<0.001) and 2 mA (*t*_*40*_=7.85, *p*<0.001) groups, but not the sham group (*t*=0.54, *p*>0.999). Between-group comparisons of PSWQ scores with adjusted p value of 0.008 showed no significant difference in the pre-intervention measurements, but significant enhancement of scores in the post-intervention measurements (1 mA vs sham: *t*_*40*_=4.52, *p*<0.001; 2 mA vs sham: *t*_*40*_=7.14, *p*<0.001; 1 mA vs 2 mA: *t*_*40*_=2.75, *p*<0.025) (Fig. 3c). This indicates that decreased worry was specific for tDCS effects with a significantly larger effect for the 2 mA group. In the BDI-II, we observed an interaction of group×time (*F*_*2,38*_=4.05, *p*=0.025, *η*p2=0.17). The main effects of group and time were significant as well (Table 3). Post *hoc* t-tests (adjusted *p*=0.016) showed a significant decrease of depressive state between pre- and post-intervention measurements in the 1 mA (*t*_*40*_=3.57, *p*=0.001) and 2 mA (*t*_*40*_=4.86, *p*<0.001) groups only. However, when compared to the sham group (adjusted *p*=0.008), a significant decrease of depressive states was observed only in the 2 mA (*t*_*40*_=5.36, *p*<0.001) but not the 1mA group (*t*_*40*_=2.44, *p*=0.056), and the magnitude of change was significantly larger in the 2 than in the 1 mA group (*t*_*40*_=3.01, *p*=0.010) (Fig. 3d). No significant differences in depressive state were found between groups in the pre-intervention measurement.

For emotion regulation abilities, a significant interaction of group×time (*F*_*2,38*_=4.16, *p*=0.023, *η*p2=0.18) was observed on the DERS score. The main effects of group and time were also significant (Table 3). Emotion regulation was significantly improved in the post-intervention compared to the pre-intervention measurement in both 1 mA and 2 mA groups (*t*_*40*_=3.62, *p*=0.001; *t*_*40*_=3.85, *p*<0.001), but not in the sham group (*t*_*40*_=0.44, *p*>0.999) (adjusted *p*=0.016). Between-group comparisons showed no significant differences in the pre-intervention measurements, but significant differences in the post-intervention measurements for the 1 mA vs sham and 2 mA vs sham conditions respectively (*t*_*40*_=3.26, *p*=0.005; *t*_*40*_=3.90, *p*=0.001), with no differences between the 1 mA vs 2 mA groups (*t*_*40*_=0.70, *p*>0.999) (Fig. 3e). Finally, the results of the 3×2×4 (quality of life factors) ANOVA revealed a significant interaction of group×time (*F*_*2,114*_=16.08, *p*<0.001, *η*p2=0.55) on the WHOQUL scores. Quality of life did not significantly interact with group and group×time indicating that all domains of quality of life were similarly affected. The main effects of group and time were, however, significant (Table 3). Post *hoc* comparisons (adjusted *p*=0.016) showed a significantly improved quality of life compound score at the post-intervention measurement in both 1 mA (*t*_*40*_=3.69, *p*<0.001) and 2 mA (*t*_*40*_=5.59, *p*<0.001) as compared to the sham stimulation group. Active tDCS groups did not significantly differ in post-intervention scores (adjusted *p*=0.008). There were no significant between-group differences in the pre-intervention measurements, but significant differences were found in the post-intervention measurements for all domains of quality of life (Fig. 3f). [Table 3 and Fig 3 here]

### 3.4. Attention bias

The results of the 2×2 ANOVA showed no significant group×time interaction or main effects of time and group on the mean bias score (Table 3). However, when we entered stimuli (threat vs neutral) in a 2×2×3 mixed ANOVA, the results showed a significant group×time interaction (*F*_*2,38*_=12.01, *p*<0.001, *η*p2=0.38) as well as significant main effects of time, group, and stimuli on the RT of task performance. Bonferroni-corrected post *hoc* t-tests (adjusted *p*=0.016) revealed a significant *pre* vs *post*-intervention RT reduction of the threat-related stimuli only in the 2 mA group (*t*_*40*_=3.12, *p*_*A*_=0.007) but not 1 mA (*t*_*40*_=1.88, *p*_*A*_=0.190) or sham (*t*_*40*_=1.39, *p*_*A*_=0.504) groups (Fig. 3g). No significant difference in the post-intervention RT was observed between the 1 mA vs 2 mA groups for both threat (*p*=0.595) and neutral stimuli (*p*=0.413). The same pattern of response was found for the neutral stimuli except that here both interventions significantly reduced RT after intervention and compared to the sham group (Fig. 3h). These results show that the intervention had an overall improving effect of patients’ RT, regardless of the stimuli valence, but did not improve the bias score in patients.

Lastly, we calculated Pearson’s correlations to see if SAD symptoms in the post-intervention and follow-up measurements correlate with attentional bias task performance. No significant correlation was found between the post-intervention bias scores and fear, avoidance and LSAS total score. We however found significant positive correlations between reduced RT for both threat-related and neutral stimuli and alleviated SAD symptoms, including *fear* symptom (threat: *p*_*post*_=0.009; *p*_*follow-up*_<0.001; neutral: *p*_*post*_<0.001; *p*_*follow-up*_<0.001), and *avoidance* symptom domains (threat: *p*_*post*_=0.009; *p*_*follow-up*_=0.006; neutral: *p*_*post*_=0.003; *p*_*follow-up*_=0.006), as well as SAD total score (threat: *p*_*post*_<0.001; *p*_*follow-up*_<0.001; neutral: *p*_*post*_<0.001; *p*_*follow-up*_<0.001).

## 4. Discussion

In this randomized, sham-controlled, parallel-group clinical trial, we investigated the impact of a novel, intensified tDCS protocol (stimulation twice per day with 20 min interval), with intensity dosage comparison (1 mA vs 2 mA) on primary symptoms, clinical measures, and attentional bias in patients with SAD. Both active stimulation conditions, compared to the sham group, significantly reduced fear and avoidance symptoms after the intervention. Fear symptom improvement continued for up to 2 months in both active groups, while avoidance was significantly reduced for up to 2 months only in the 2 mA stimulation group. In treatment-related measures, except for depressive state, which was significantly reduced only in the 2 mA group, other measures (worries, emotion regulation, quality of life) were significantly improved in both active stimulation conditions versus the sham group after the intervention. Moreover, depressive symptoms and worries reduction were significantly larger in the 2 mA vs the 1 mA group. Finally, we found that only the 2 mA protocol reduced RT of both threat-related and neutral stimuli as compared to the sham group although attention bias score was not significantly different across groups.

Reduction of SAD symptoms and clinical improvement following active stimulation can be first and foremost explained via the assumed changes in the lateral-medial prefrontal network, which is involved in the pathophysiology of SAD. Our findings support the assumed pathophysiological mechanism that impaired executive control network functionality (involving DLPFC), along with disturbed motivational and emotional networks (involving VMPFC, amygdala) are central for the development and maintenance of SAD symptoms[18, 20]. The stimulation protocol in our study was designed based on evidence showing a hypoactivation of the lateral PFC[58, 59] along with exaggerated activation of the medial PFC in trait anxiety, SAD[14, 18, 21] and anxiety disorders[10, 13] although functional abnormalities have not been consistent across studies. Based on these studies, we speculate that anodal stimulation-generated excitability enhancement of the DLPFC increased functional connectivity of this region with SAD-relevant networks hereby increased control of the threatening stimuli. On the other hand, cathodal stimulation of the medial PFC was expected to decrease excitability of this region and modulate its connectivity with emotion networks and hereby alters emotional experience of social threats. Reduced *fear* induced by, and *avoidance* of social stimuli, lower worries and depressive states, and improved emotion regulation may be explained via concurrent upregulation of executive control and downregulation of threat-sensitivity (Fig. 4). Indeed, such cognitive control-emotion regulation association and its improvement via prefrontal tDCS has been shown in other neuropsychiatric disorders associated by emotional disturbances[60-62].

**Fig 4.**
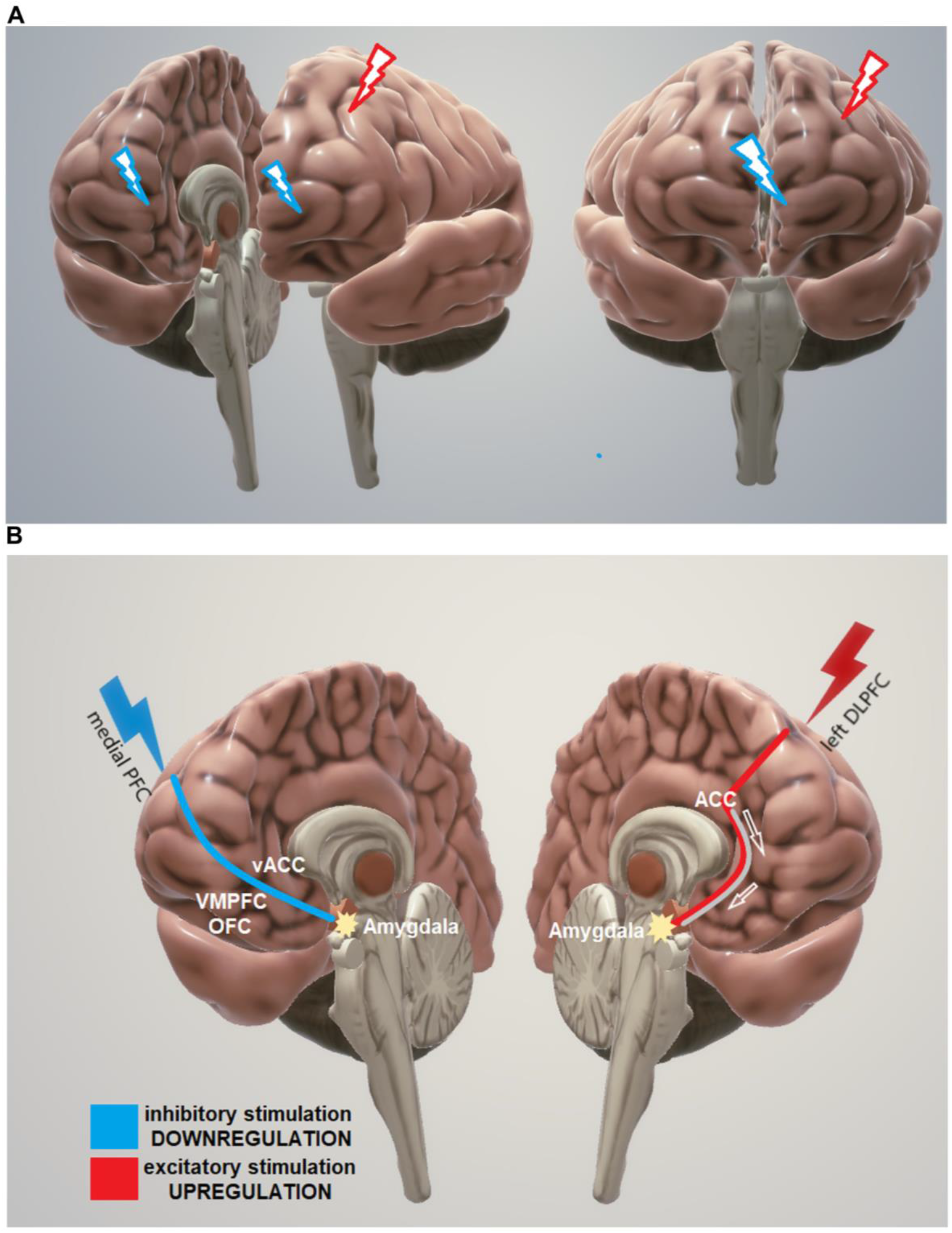
Expected regulatory effects of anodal left DLPFC-cathodal medial PFC on SAD symptoms and attentional bias to threat. (**A**) Expected regulatory effects of anodal left DLPFC-cathodal medial PFC on SAD symptoms and attentional bias to threat. Anterior and medial-lateral view of stimulation targets with anodal (red) and cathodal stimulation (blue). (**B**) Excitatory stimulation of the left DLPFC and nearby regions (e.g. ACC) is expected to upregulate attentional and executive control over the threat, anxiety-provoking stimuli, and negative emotions, thereby indirectly modulating amygdala hypersensitivity. Inhibitory stimulation of the medial PFC is expected to reduce the activity of these regions (VMPFC, medial OFC) and the amygdala, thereby downregulating negative emotions and overreactions to threatening stimuli. It is of note, this is a simple proposed mechanism for the effects of the applied intervention (anodal left DLPFC, cathodal medial PFC). While this model considers mostly the cognitive control network (related to DLPFC) and motivational network related to medial PFC, ventral ACC, and subcortical regions, other involved networks in SAD including the default mode network (DMN) and dorsal attention network (DAN) are not depicted in this model. *Note*: PFC = prefrontal cortex; DLPFC = dorsolateral prefrontal cortex; ACC = anterior cingulate cortex; vACC = ventral ACC; VMPFC = ventromedial prefrontal cortex; OFC = orbitofrontal cortex.

Moreover, we found overall reduced attention to both threat-related and neutral stimuli which were specific to the 2 mA group. Although we expected a specific reduced bias to threat-related stimuli only, this can be interpreted as an overall enhancement of attentional resources, regardless of the valence of stimuli as a result of DLPFC activation. In accordance, in healthy individuals anodal left DLPFC stimulation (2 mA) reduced vigilance to threatening stimuli in a similar dot-probe task[27]. In another tDCS-fMRI study in individuals with high trait anxiety, active DLPFC stimulation increased activity of cortical regions associated with attentional control and was associated with behavioural improvement in a threat-related attentional task[63]. Our findings did not show a threat-specific reduction of attention bias, but an overall performance enhancement (i.e., reduced RT) for all stimuli which can be attributed to the facilitation of attentional resources as a result of prefrontal tDCS[64, 65].

In addition to the cortical regions (DLPFC, medial PFC) which are assumed to be directly modulated by our intervention, there are subcortical regions responsible for assigning salience to threatening stimuli, which are connected to the deficient cognitive control and motivation networks (i.e., lateral-medial PFC). Lateral and medial PFC regions are connected with subcortical areas like the insula and amygdala[66, 67]. Previous studies have shown that activation of the DLPFC with anodal tDCS modulates reactivity of the insula[62] and amygdala[63] to threat, regions that are hyperresponsive to threat and thus involved in the pathophysiology of SAD[21]. Interestingly, it has been shown that effective amygdala-prefrontal connectivity predicts successful emotion regulation[68], the variable which significantly improved in both active tDCS groups and is linked to SAD core symptoms (fear, avoidance). Accordingly, we assume that our stimulation protocol affected also the emotional network, which is hyper-responsive to social situations in SAD, and linked with cognitive control and motivation networks[18]. It is important to consider here that involved networks in SAD, which also include the parietal-occipital networks, cannot be reduced to prefrontal-cingular network and thus other areas might be promising candidates for brain stimulation interventions in SAD.

A main objective of this study was to compare the efficacy of an intensified stimulation protocol (twice per day with 20 min interval, as compared to the more conventional once per day protocols), delivered with different intensities (1 vs 2 mA). This protocol has not been explored before in SAD and other anxiety disorders to the best of our knowledge. The rationale behind the protocol comes from a study showing that twice stimulation with 20 min interval leads to longer aftereffects on cortical excitability compared to non-repeated stimulation or stimulation with long intervals, and resembles features of late-phase LTP,[43, 69] as well as a recent study demonstrating that 20 min of anodal stimulation-induced larger aftereffects for both 1 and 2 mA intensities, as compared to other durations (15 and 30 min)[70]. We chose specifically 1 and 2 mA stimulation intensities because results of previous studies show that the effects of stimulation on psychological processes are not in all cases larger with higher intensity[71, 72]. Our findings showed that overall, both interventions in this intensified protocol significantly reduced SAD symptoms, worries, depressive states and emotion regulation after intervention. However, the 2 mA stimulation condition led to significantly larger effects as compared to the 1 mA condition for some measures including depressive states and worries after intervention, and the avoidance symptom at both post-intervention and 2-month follow-up measurements (Fig.3). Moreover, attentional bias to the threat-related stimuli was significantly reduced only after 2 mA stimulation, while 1 mA protocol induced only a trend-wise change.

These results have at least three clinical implications. First, intensified stimulation (twice per day with a 20 min interval), has significant acute clinical efficacy for SAD symptoms and treatment-related variables. This is in line with physiological studies that have shown that repeated tDCS sessions induce larger increases of excitability,[73] as well as with the results of clinical studies in which larger efficacy with repeated sessions was reported[38]. Here, stimulation duration (e.g. 20 min) and the interval between stimulations (e.g. 20 min) are also important as shown in previous studies which were considered in our protocol[43, 69, 70]. Second, 2 mA stimulation is associated with higher clinical efficacy in SAD, and probably in other anxiety disorders. This is a novel finding with important clinical implications. Although some other studies reported no outcome differences between 2 mA vs 1 mA tDCS in healthy individuals[70, 73], our study showed a relatively larger clinical efficacy induced by 2 mA tDCS with this specific intensified protocol. Partially heterogeneous effects between studies might be based on different factors, including different neurotransmitter availability and electrode distance to the brain in respective cortical regions under study (e.g. motor vs prefrontal). Finally, it should be noted that because we could not secure follow-up measures on most outcome parameters, the long-term clinical efficacy of this intervention cannot be evaluated with certainty by this preliminary, but promising data.

It is also important to interpret the observed effects, which are remarkably large, with some precautions due to specific limitations. First, follow-up measures could not be secured for all measures (due to pandemic restrictions) except for the LSAS scores. Therefore, magnitude and duration of the treatment efficacy should be the topic of future replication studies. Small sample size is another detrimental factor in interpreting the results. Finally, neurophysiological and brain functional measures (e.g., fMRI, EEG, TMS-EEG) were not obtained in the present study, but would be valuable to provide a comprehensive picture of treatment efficacy, and clarify mechanisms. This is important, because all of the measures used in this study, except for the dot-probe task, were based on subjective self-reports scales. Nevertheless, a potential placebo effect is unlikely as the effects were not observed in the sham group.

Taken together, our findings suggest that the intensified prefrontal tDCS condition introduced for the treatment of SAD in the present study is promising. Both, primary SAD symptoms (fear, avoidance) and secondary treatment-related variables (worries, depressive state, emotion regulation, quality of life) improved after the intervention (compared to baseline performance). Selective attention to both threat-related and neutral stimuli was significantly reduced after the 2 mA intervention as well. 2 mA tDCS, as compared to 1 mA tDCS, appears to induce larger effects on some clinical and behavioral performance measures. The observed effect sizes, however, are unusually large compared to previous tDCS works. Considering the relatively small sample size and variability in expression of anxiety, our results need to be replicated in larger clinical trials addressing the following limitations. First, our follow-up measures were compromised due to the current pandemic and we could only remotely measure SAD symptoms. Long-term improvement of other measures including attentional bias to threat was not investigated. Furthermore, the intrinsically limited focality of tDCS can result in a relatively diffuse stimulation, and thus additional cortical and subcortical areas might also have been affected. Neuroimaging methods will help to identify the regions directly affected by tDCS more accurately in future studies. Future studies need to examine the magnitude and duration of effects in studies with longer follow-up assessments and added physiological measures.

## Data Availability

The data that support the findings of this study are openly available at 10.17605/OSF.IO/2TKB3

## Acknowledgments

MAN receives funding by Deutsche Forschungsgemeinschaft (DFG – Project number 316803389) and by the German Federal Ministry of Education and Research (BMBF - GCBS grant 01EE1501), Germany.

## CRediT authorship contribution statement

**Eisa Jafari & Jaber Alizadehgoradel**: Resources, Project administration, Investigation, Data curation, Validation. **EJ** takes responsibility for the integrity and accuracy of the data. **Fereshteh Pourmohseni Koluri** & **Ezzatollah Nikoozadehkordmirza:** Resources, Project administration. **Meysam Refahi** & **Mina Taherifard**: Investigation, Data curation. **Vahid Nejati**: Software, Resources. **Amir-Homayoun Hallajian**: 3D Modeling. **Elham Ghanavti**: Writing - review & editing, Visualization, Modeling. **Carmelo M. Vicario**: Supervision, Writing - review & editing. **Michael A. Nitsche**: Supervision, Methodology, Writing - review & editing. **Mohammad Ali Salehinejad**: Conceptualization, Methodology, Supervision, Writing – original draft, Writing - review & editing, Formal analysis. All authors read and approved the final version of the manuscript.

## Data availability

The data that support the findings of this study are openly available at 10.17605/OSF.IO/2TKB3

### Abbreviations

SAD: Social anxiety disorder
DLPFC: dorsolateral prefrontal cortex
tDCS: transcranial direct current stimulation

